# Using mobile health technology to assess childhood autism in low-resource community settings in India : an innovation to address the detection gap

**DOI:** 10.1101/2021.06.24.21259235

**Authors:** Indu Dubey, Rahul Bishain, Jayashree Dasgupta, Supriya Bhavnani, Matthew K. Belmonte, Teodora Gliga, Debarati Mukherjee, Georgia Lockwood Estrin, Mark H. Johnson, Sharat Chandran, Vikram Patel, Sheffali Gulati, Gauri Divan, Bhismadev Chakrabarti

## Abstract

Autism Spectrum Disorders, hereafter referred to as autism, emerge early and persist throughout life, contributing significantly to global years lived with disability. Typically, an autism diagnosis depends on clinical assessments by highly trained professionals. This high resource demand poses a challenge in resource-limited areas where skilled personnel are scarce and awareness of neurodevelopmental disorder symptoms is low. We have developed and tested a novel app, START, that can be administered by non-specialists to assess several domains of the autistic phenotype (social, sensory, motor functioning) through direct observation and parent report. N=131 children (2-7 years old; 48 autistic, 43 intellectually disabled, and 40 typically developing) from low-resource settings in the Delhi-NCR region, India were assessed using START in home settings by non-specialist health workers. We observed a consistent pattern of differences between typically and atypically developing children in all three domains assessed. The two groups of children with neurodevelopmental disorders manifested lower social preference, higher sensory sensitivity, and lower fine-motor accuracy compared to their typically developing counterparts. Parent-report further distinguished autistic from non-autistic children. Machine-learning analysis combining all START-derived measures demonstrated 78% classification accuracy for the three groups (ASD, ID, TD). Qualitative analysis of the interviews with health workers and families (N= 15) of the participants suggest high acceptability and feasibility of the app. These results provide proof of principle for START, and demonstrate the potential of a scalable, mobile tool for assessing neurodevelopmental disorders in low-resource settings.

## INTRODUCTION

Autism Spectrum Disorder (ASD) is an early-onset neurodevelopmental condition with a global prevalence of ∼1% and is a priority for the global mental health agenda (Baxter et al., 2015). It is estimated that India is home to ∼5 million families with a child with ASD^1^ (Arora et al., 2018). In India, as in many low-resource settings, most families do not receive any, let alone timely, intervention, in part because most never receive a diagnosis (Durkin et al., 2015; Krishnamurthy, 2008; Samms-Vaughan, 2014). Even those who are diagnosed experience significant delays between parents’ noticing differences in behaviour and clinical diagnosis by professionals (Jain et al., 2013; Samms-Vaughan, 2014). Low community awareness about ASD leads to reduced help-seeking behaviour (Minhas et al., 2015), and is exacerbated by a number of other challenges to detection. First, there is a paucity of skilled human resources serving diagnostic services to a population of over 1.2 billion such as developmental practitioners, psychiatrists, neurologists, and psychologists (Kumar, 2011). When available, these specialists work in urban areas inaccessible to the large rural population. Second, current diagnostic approaches typically involve time-intensive, expensive and proprietary tools, greatly limiting access (Durkin et al., 2015). Most of these tools are either not adapted to local cultures or not available in local languages (Soto et al., 2015). Third, social stigma prevents parents’ seeking a psychiatric diagnosis for their child (Minhas et al., 2015).

Yet there is emerging evidence from low- and middle-income country settings that non-specialist health-worker delivered, parent-mediated intervention targeting social communication is acceptable and effective in improving outcomes for autistic children (Rahman et al., 2016), consistent with similar evidence from high-income countries (Oono et al., 2012). In light of such evidence that task-shifting, community-based interventions can close the treatment gap, the detection gap becomes an urgent priority, highlighting the need for proactive screening for ASD. The current study aimed at developing a tool that could be used by non-specialists to assess ASD risk in low-resource settings, allowing the closing of the detection gap.

Mobile technologies offer a significant advantage in this effort, given their wide penetration across geographies and socioeconomic strata. The increasing availability and affordability of these devices, coupled with the steady advances in sensor technology and portable processing power, makes these technologies ideally suited for scalability. Parallel efforts to harness mobile technology to assess autism-related behavioural phenotypes have shown promise in high-resource settings (Dawson & Sapiro, 2019; Egger et al., 2018). In the current study, we aim to test a battery of tasks that index different aspects of the autistic phenotype, using a mobile device. In view of the diverse phenotypic domains associated with ASD, the mobile platform (app) includes direct assessments of the child on multiple tasks that relate to social behaviour, sensory interests, and motor function. The platform also includes assessment of parent-reported autistic features through a questionnaire, and an observational measure of parent-child interaction.

This validation exercise aims to test whether this assessment can distinguish children with neurodevelopmental disorders, and in particular autism, from their typically developing counterparts. To this end, we have implemented and benchmarked the assessment in the form of a scalable, mobile tool, administered in the community by non-specialists to assess autism-related features in 2-to-7-year-old children in home settings in India.

## METHODS

### Participants

Three groups of children were recruited: (1) children with a diagnosis of Autism Spectrum Disorder (ASD), N=48; (2) children with a diagnosis of Intellectual Disabilities (ID), N= 43; and (3) typically developing (TD) children N=40 (see Table 1). The ASD and ID groups were recruited by contacting the families of children attending outpatient paediatric clinics at a tertiary clinic (All India Institute of Medical Sciences, Delhi, India). The TD group was recruited through snowballing from communities from where children with ASD and ID were recruited. - The three groups were matched for chronological age. The ASD and ID groups were matched on cognitive age using a language-adapted version of the *Developmental Profile-3 (DP3)* (Alpern, 2007). The ASD group was compared with the other two groups for the severity of autistic symptoms using a locally developed and standardised tool, the *INCLEN Diagnostic Tool for Autism Spectrum Disorder (INDT-ASD)* validated against DSM-V criteria (Juneja et al., 2014).

**Table 1:** Participant characteristics

| | TD | ASD | ID | F/$\chi^2$ | p-value | Post-hoc contrasts,<br>p-value |
| --- | --- | --- | --- | --- | --- | --- |
| **Chronological Age**<br>**M ± SD** | (N=40)<br>4.59 ± 1.34 | (N=48)<br>4.24<br>± 1.22 | (N=43)<br>4.56<br>± 1.67 | F (2, 129) = 0.88 | 0.42 | |
| **Gender ratio**<br>**(F:M)** | 19:21 | 12:36 | 9:34 | 7.99 | 0.02 |  |
| **Cognitive age on DP3** | (N=36)<br>4.32<br>± 1.49 | (N=37)<br>1.49<br>± 0.53 | (N=36)<br>1.94<br>± 0.80 | F (2, 106) = 80.87 | <0.0001 | TD > ASD, <0.0001<br>TD > ID, <0.0001<br>ID ~ ASD, 0.19 |
| **INDT-ASD** | (N=37)<br>0.16 ± 0.37 | (N=37)<br>17.16 ± 4.35 | (N=39)<br>5.15 ± 7.51 | F (2, 110) = 109.97 | <0.0001 | TD < ASD, <0.0001<br>TD < ID, <0.0001<br>ID < ASD, <0.0001 |

### Tools

The START task battery was administered on all participants alongside standardized tools of autism symptom severity and developmental level.

#### Screening Tool for Autism Risk using Technology (START) task battery

START is an Android app presented on a mobile device, that can be administered by non-specialists with minimal training. The app includes a battery of tasks measuring multiple domains of development (Table 2). This choice of domains was informed by the developmental differences commonly identified in children with autism.

Differences in *social* behaviour are a core diagnostic feature of autism. Lab-based experiments designed to measure this aspect of the autistic phenotype have often focussed on presenting social alongside nonsocial stimuli (Dubey et al., 2015; Pierce et al., 2011; Ruta et al., 2017). Such paradigms have revealed that autistic individuals have reduced preference for social stimuli, and make less effort to seek out social over nonsocial stimuli (Hedger et al., 2020). Accordingly, the START task battery includes two measures of social reward responsivity: 1) a passive viewing paradigm similar to the eye-tracking laboratory-based task of Pierce et al. (2011), and 2) a choice-based paradigm similar to that of Ruta et al. (2017).

Atypical *sensory* sensitivity is a commonly reported feature of autism (Ausderau et al., 2014; Ben-Sasson et al., 2007; Posar & Visconti, 2018). It is generally evaluated using parent-report questionnaires or tasks that involve touching/watching objects of special sensory interest (e.g. spinning wheels with illusory contours, pin cushions, musical dome). The START task battery includes an adapted version of one such lab-based task used by Tavassoli et al. (2016) to measure visual sensory sensitivity.

Atypical *motor* skills are commonly reported in autism (Anzulewicz et al., 2016; Ghaziuddin et al., 1994; Manjiviona & Prior, 1995). Poor spatial coordination and weak adaptation of velocity to reach targets have been suggested to be specific to autism (Forti et al., 2011). Developments in touch sensor technology can help measure spatial coordination and velocity with high precision and ease. The START task battery harnesses this technological development to measure three-dimensional finger movements, providing a fine-grained measure of spatio-temporal performance in fine-motor planning and execution.

Behavioural observations may emerge from parent reports of day-to-day activities of the child, or expert observation of social interaction and play. Brief parent-report tools such as the INCLEN Diagnostic Tool for Autism Spectrum Disorder (INDT-ASD) (Juneja et al., 2014), and All India Institute of Medical Sciences (AIIMS)-Modified-INDT-ASD Tool (Gulati et al., 2019) have demonstrated high sensitivity in early screening and diagnosis of ASD in an Indian setting. Accordingly, the START app includes a brief questionnaire for primary caregivers as well as a provision for video-recording a parent/caregiver-child play session. Dyadic interaction of the child with the caregiver constitutes one of the most ecologically valid metrics of social interaction, and is the primary target of certain types of developmental interventions for autism (Green et al., 2010).

#### Standardised tools to evaluate level of functioning and autism symptom severity

The *Developmental Profile 3 (DP3)* (Alpern, 2007) is a parental interview scale designed to assess development and functioning across five areas: physical, adaptive, social-emotional, cognitive and communicative. We used the age-equivalent score from the cognitive subscale to estimate development that is not influenced by specific difficulties in social or communicative function. The *INCLEN Diagnostic Tool for Autism Spectrum Disorder (INDT-ASD)* (Juneja et al., 2014) is specifically developed for diagnosing autism in 2-9-year-old children in India. It has a high validity against DSM-IV-TR diagnoses and Childhood Autism Rating Scale (Schopler et al., 1980) scores as well as with DSM-V (Vats et al., 2018).

These tools were administered by trained postgraduate psychologists.

#### Evaluation of acceptability and feasibility of the START tool

Research assistants (1) completed a detailed observation schedule noting the environment and circumstances of each data collection, including family involvement and available resources, (2) interviewed non-specialist health workers both immediately after their training and at the end of data collection, with a focus on challenges faced during data collection and strategies adopted to overcome these, and (3) interviewed parents of participating children (TD=5, ASD=5, ID=5) to explore their experiences with START, including caregivers of children who were able to complete the START assessment tasks and those who were unable to complete them. Separate consent for audio recording was taken prior to these interviews. Further details of the observation and interview schedules are available in the supplementary material.

### Procedure

Two high school graduates with no prior experience in mental health or child assessments or in using tablet computers, were recruited as non-specialist health workers. They were provided a structured two-day classroom training followed by two-day observation and supervised field training in households. Classroom training included START app setup and administration using a detailed statement of procedure. Supervised training with TD child volunteers in the classroom was followed by field training wherein they observed a researcher administer START on one TD and one atypically developing child. They then practised the data collection on two TD volunteers.

Once trained, a health worker and a research assistant visited participants’ households to collect data. The health worker administered START on a Samsung SM P600 tablet while the research assistant took field notes, monitored quality of data collection and helped ensure a conducive testing environment. Health workers used available resources within the households to create the best possible testing environment, particularly around seating and lighting. Testing was generally conducted sitting on the floor or bed.

## Data analysis

Pre-set exclusion criteria were applied to the data to ensure quality. This resulted in a different number of participants’ data for each task. Detailed information about the criteria used for each task, the final N for each task, and the data analysis is given below.

### Preferential Looking Task

Gaze location was identified using a convolutional neural net-based algorithm, based on the gaze-capture algorithm (Bishain, 2019; Dubey, Brett, Ruta, Bishain, Chandran, Bhavnani, Belmonte, Estrin, Johnson, Gliga, & Chakrabarti, 2019; Krafka et al., 2016). Data were collected from 118 of 131 participants (TD = 40, ASD=40, ID=38). All participants met the inclusion criteria of eye detection for at least 50% of frames as well as gaze on the tablet for at least 50% of frames. Social preference was computed as a ratio between the number of frames during which a participant was gazing at the social stimulus and the total number of frames in which their gaze was identified to be on either of the two stimuli.

### Button Task

Data were collected from 116 of 131 participants (TD = 40; ASD = 37; ID = 39). Participants who completed fewer than 50% of trials were excluded from the analysis. A trial in this context refers to an instance of the two buttons being presented (Table 2). This exclusion criterion yielded data from 104 participants (TD=39; ASD=27; ID=38) in the final analysis. For each participant, the proportion of social button choice as a fraction of the total number of completed trials was calculated.

### Wheel Task

Data were collected from 125 of 131 participants (TD = 40, ASD=46, ID=39). Data were filtered for quality by removing participants who completed fewer than two of the five trials, or whose face could be detected in only 25% or fewer of the video frames. A trial was considered complete if it was not aborted. This exclusion criterion yielded data from 117 of 131 participants (TD = 37, ASD=41, ID=39) in the final analysis. Two variables were coded from this data set: a) Time spent looking at the wheel, and b) distance of the face from the screen. The choice of these variables was based on the observations that autistic children tend to watch the spinning wheel for longer and come closer to it than their TD counterparts (Tavassoli et al., 2016). Time spent looking at the wheel was calculated for every completed trial, summed across trials and divided by the maximum possible duration of the completed trials. Distance of the face from the screen was calculated using a deep neural network that detected the subject’s facial features in each frame (Bishain et al., 2020). Distance of the head to screen has previously been shown to be related to interest (Toker et al., 2017; Witchel et al., 2020).

### Motor Following Task

Data were collected from 120 of 131 participants (TD = 40, ASD=43, ID=37). Data sets were filtered for completeness by including only participants who completed two or more trials. This criterion yielded 115 participants (TD = 40, ASD=40, ID=35) for the final analysis. Spatio-temporal difference between the target and the child’s motor trajectory was computed as root mean square error (RMSE) to measure accuracy in motor planning and execution. This measure utilized the error in the simultaneous positions of the butterfly and child’s touch point. Additionally, we analyse the ‘frequency gain’ metric for all participants using a Fast Fourier Transformation (FFT). The trajectories of the butterfly and child’s trace were resolved into multiple waves of varying amplitudes using FFT. This allowed us to analyze the closeness in the source and target motions along the frequency domain (for details see Supplementary Material).

### Bubble Popping Task

Data were collected from 120 of 131 participants (TD = 40, ASD=41, ID=39). Data were included from all the participants who popped one or more balloons. Force used while popping the balloons was recorded using the getPressure parameter recorded by the Android operating system on Samsung tablets, and averaged across all balloons popped. Distance between the touch point and the centre of the balloon was calculated to estimate visuomotor targeting accuracy in approaching dynamic stimuli.

### Colouring Task

Data were collected from 113 of 131 participants (TD = 40, ASD=38, ID=35). Participants were asked to colour the interior of a target figure. Data sets were included only if participants coloured at least 25% of the pixels on the screen. This criterion yielded 93 participants (TD = 37, ASD=29, ID=27) in the analysis. The total number of crossings over the target figure’s outlines (movements in and out of the figure) was calculated. Any change in the touch point from inside the figure (pixels identified inside the outline) to outside or *vice versa* was counted as one crossover.

### Parent/Caregiver- Child Interaction

Data were included from 100 of 131 participants (TD = 32; ASD = 35, ID = 33). The video-recording of the session was coded using the Dyadic Communication Measure for Autism, a standardised framework for coding dyadic interaction between parents and children by three independent coders (Green et al., 2010). Two measures were extracted from this data set, one indexing the child’s attempts at initiating interactions, and the other indexing synchronous responses from the caregiver. Child initiations were scored as a percentage of the total number of attempts to interact with the caregiver, which included a) initiating interactions, b) responding to caregiver, and c) other behaviours. Similarly, caregiver responses were scored as the percentage of synchronous interactions within the total number of interactions. 13% of the videos were coded by all the three coders and used to calculate intra-class correlation (ICC) using a 2-way mixed-effects model, based on single measure, absolute agreement and confidence interval of 95%. A high degree of reliability was found between the coders for scores on parent/caregiver’s synchronous interaction as ICC was 0·876 (*p <*.*0001*, 95% CI [.69, 96]). However the coders had limited reliability for the scores on child’s initiation as ICC was .542 (*p <*.*0001*, 95% CI [-.04, .85]). Where the videos were coded by more than one coder, we randomly chose codes from any one coder.

### START-ASD Questionnaire

Data were collected from all 131 participants (TD = 40, ASD =48, ID =43). The items were scored as binary responses. The summed score indicates the number of ‘red flag’ signs of ASD.

For each task, the three groups were contrasted on the dependent variables defined above using analyses of variance (Table 3). The Kruskal-Wallis test was used where the assumption of normality was violated; and Welch and Brown-Forsyth robust tests were run where the assumption of homogeneity of variance was violated. Since the results from these alternative analyses were similar to those obtained with the general linear model, we report in Table 3 results from the standard analysis of variance. Results from the alternative statistical tests are presented in Supplementary Tables 2 and 3.

### Machine-learning analysis

This analysis applied a data-driven technique to combine the information from the several measures within START so as to optimise discrimination between the three groups (ASD, ID, TD). In this analysis, each dependent variable from the individual tasks constituted a feature vector. These features were then subjected to a set of machine-learning methods including XGBoost, logistic regression, and support vector machines. First, each feature was evaluated independently for its accuracy in classifying individuals into the three groups. In addition, all possible (non-singleton) combinations of the features were explored to determine a set of features optimised for classification. This combinatorial exploration identifies a suite of tasks that yields the most accurate classification (see Supplementary material 1.2 for details).

### Acceptability and feasibility

Interviews were conducted with the non-specialist health workers and caregivers to evaluate feasibility and acceptability of the START task battery in home settings.

Environmental conditions for data capture and nature and frequency of disruptions during the assessment were assessed from the observation schedule used by the research assistant. All interviews were audio-recorded, transcribed and then translated to English and cross-checked for accuracy of translation. In-depth interviews were qualitatively analysed using thematic analysis. Researchers JD and DV independently familiarised themselves with the data by re-reading transcripts, following which the emerging codes were highlighted. Codes were then organised into themes and subthemes using NVIVO. Data were synthesised and triangulated using an iterative process of consensus validation between researchers.

## RESULTS

Results are presented below in three sections: a) group comparisons in the START measures, b) group classification accuracy using machine-learning analysis, and c) feasibility and acceptability of START administration by non-specialists in households.

### a) Group comparisons

We examined group differences in social, sensory, motor functions, parent/caregiver report, and dyadic interaction. For each of these domains, the three groups were contrasted on the stated dependent variables (Table 3). In the social domain, an effect of group membership is seen on the preferential looking task, as children with neurodevelopmental disorders (ASD and ID) looked at the social stimuli less than the TD group did. However, no such group difference was seen in the active choice task (button task). In the sensory domain, an effect of group membership was noted in time looking at the spinning wheel: Children with ASD and ID looked at the spinning wheel longer than their TD counterparts did. In the motor domain, neurodevelopmentally disordered groups were distinguished from TD by force in the bubble-popping task and by visuomotor accuracy across all the motor tasks. Finally, an effect of group membership was found in measures of parent/ caregiver-report and interaction. Parents of children with ASD endorsed higher numbers of items from the START-ASD questionnaire than parents of either ID or TD children. An overall inspection of Table 3 suggests a consistent pattern of group difference for neurodevelopmental disorder and TD groups. However the two neurodevelopmental disorder groups (ASD and ID) are not clearly discriminable from most of the measures (the questionnaire data and visuomotor RMS error being notable exceptions).

### b) Machine-learning analysis

The classification accuracies, the sample sizes for each group and other details as determined in the machine-learning analysis are provided inSupplementary Table 1. Based on these results, the Motor task (RMSE in following the trajectory) was the most promising independent task with 60% overall classification accuracy into three groups (TD, ID, ASD), superior to a random chance classification accuracy of approximately 33%. This discrimination accuracy is at par with that reported by the questionnaire measure (Figure 4).

**Figure 1:**
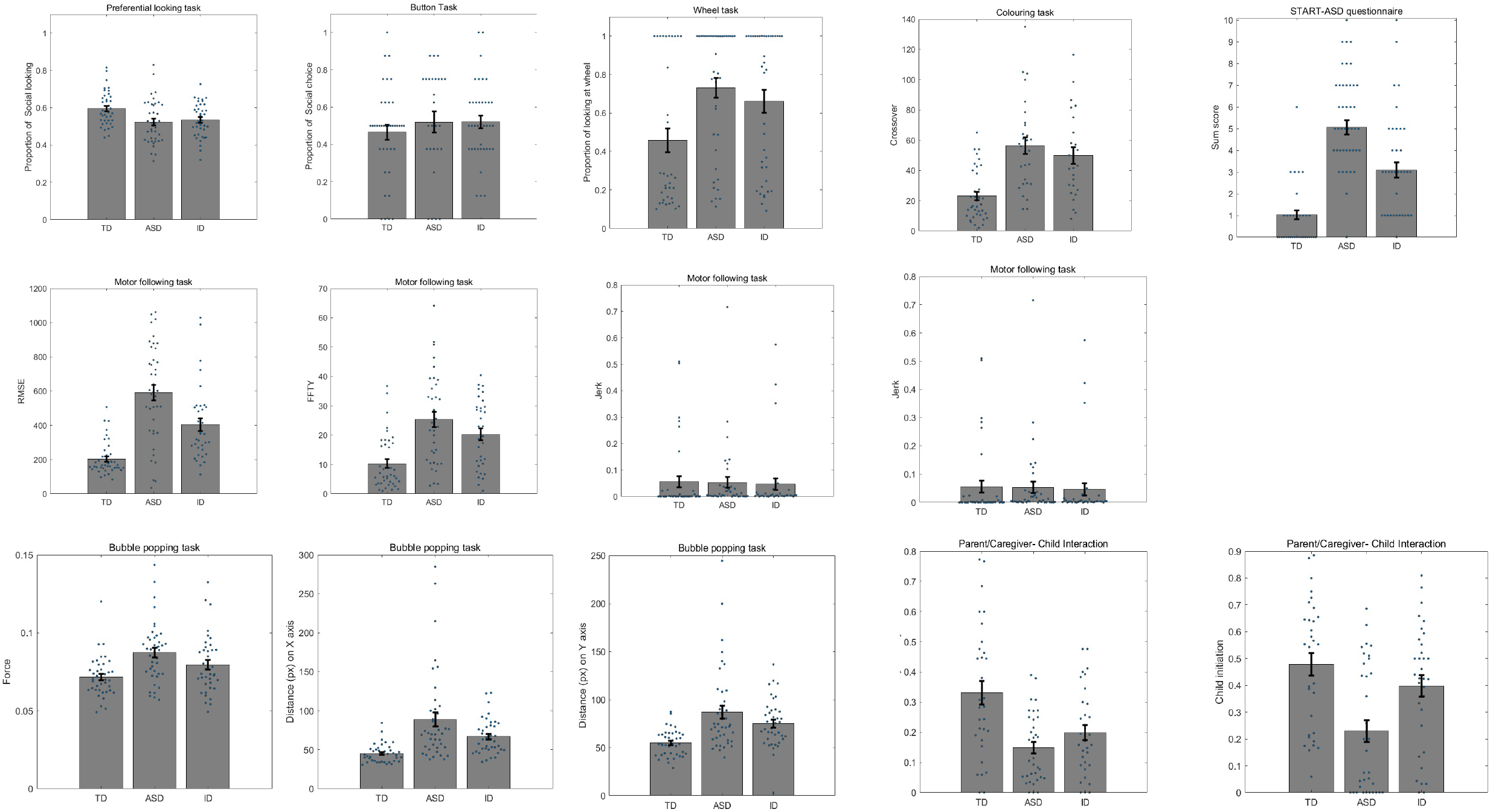
Administration of the START task battery in field settings. a) Tablet positioned upright for preferential looking task, and wheel task; b)Tablet positioned flat on a surface with a frame underneath for the button task, motor following task, bubble popping task, and colouring task. Note: Figures 1a and 1b have been removed from this preprint due to the policies of the medrxiv repository, and are available from the corresponding author upon request.

**Figure 2:**
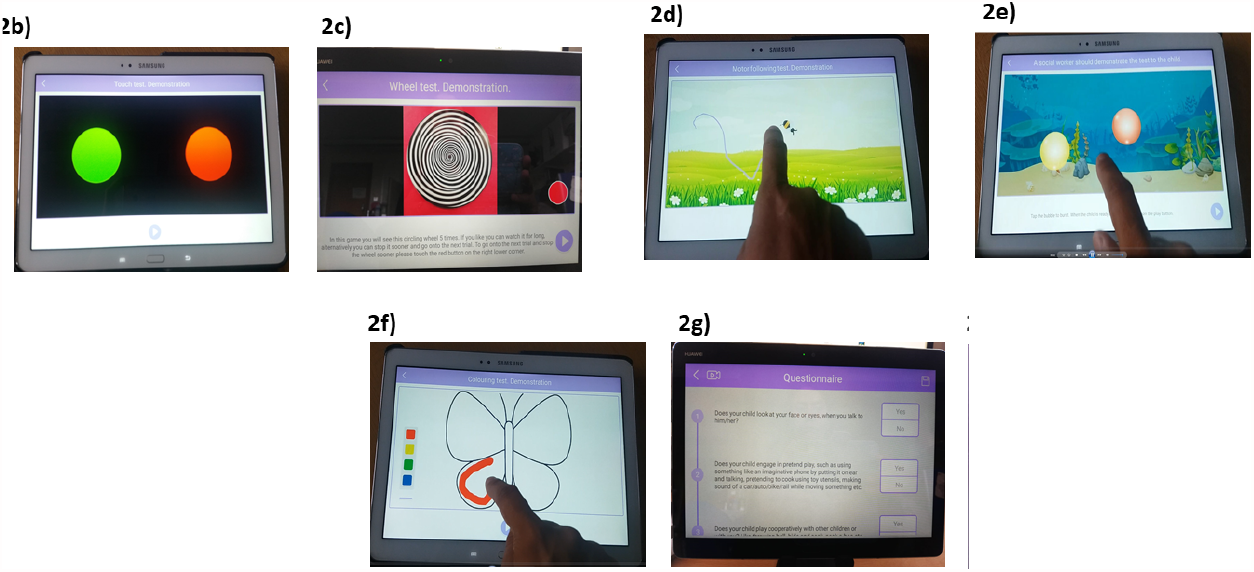
Sample screenshots from the a) preferential looking task, b) button task, c) wheel task, d) motor following task, e) bubble popping task, f) colouring task, g) START-ASD questionnaire, and h) caregiver-child interaction observation. Note: Figures 2a and 2h have been removed from this preprint due to the policies of the medrxiv repository, and are available from the corresponding author upon request.

**Figure 4:**
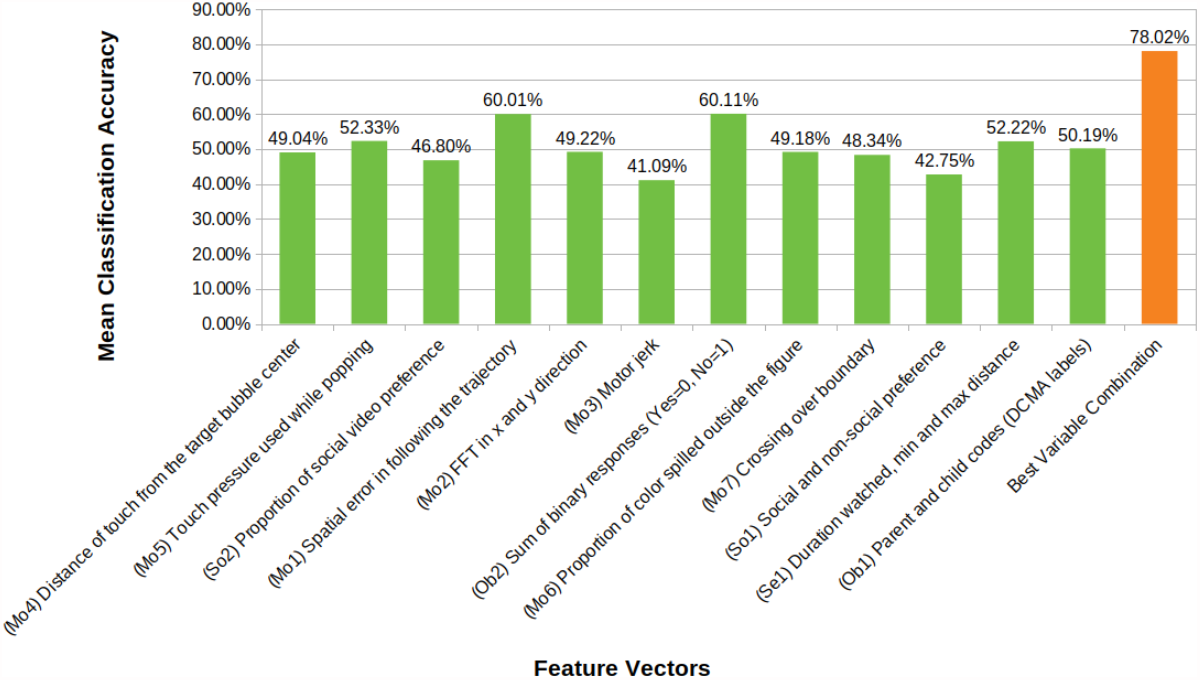
Bar graphs showing comparison of mean classification accuracies of the feature vectors taken from the eight tasks included in the START battery. The figure also represents the best classification accuracy achieved by a combination of these features. (Prefixes on x-axis in brackets refer to corresponding feature IDs). Note that some features are designed as multidimensional vectors with different measures along its dimensions

While the overall classification accuracy for each of the tasks is relatively weak, we observe a significant improvement when various combinations of the corresponding features are analysed together. The predictive power of the weaker features is significantly boosted and we report a much stronger overall classification accuracy of 78% for the three groups of ASD, ID, and TD (See Table 4). The final set of features comprised three motor / visuomotor features from three tasks-RMS error in visuomotor following, boundary crossings in colouring, and force in bubble-popping; time watched and variation in distance from the display in the wheel task; both gaze and choice measures of social preference; and video-coded and questionnaire measures of autistic behaviour.

**Table 4:** Machine learning results. The overall classification accuracy for the best combination of feature vectors is listed. Refer to Figure 4 for corresponding Feature Vector IDs. So1: Button Task, So2: Preferential Looking task, Se1:Wheel task, Mo1: Motor Following Task, Mo5, Mo7: Bubble-popping task, Ob1: Parent Child Interaction, Ob2: Questionnaire responses

| Feature Vector ID<br>combination providing<br>the best accuracy | Mean<br>Classification<br>Accuracy<br>(ASD) | Mean<br>Classification<br>Accuracy<br>(ID) | Mean<br>Classification<br>Accuracy<br>(TD) | Mean<br>Overall<br>Classification<br>Accuracy | Mean proportion %<br>of subjects across<br>different groups<br>(ASD:ID:TD) |
| --- | --- | --- | --- | --- | --- |
| **Social:** So1, So2<br>+<br>**Sensory:** Se1<br>+<br>**Motor:** Mo1, Mo5, Mo7<br>+<br>**Observation:** Ob1, Ob2 | 61.61% | 78.23% | 86.40% | 78.02% | 23:30:47 |

### c) Feasibility and Acceptability

High completion rates (>70%) were obtained in all task measures collected (see Figure 5). The two main drivers behind missing data were a) children’s unwillingness to play a game, seen more often in atypical children compared to typically developing ones and b) app malfunctions for specific tasks.

**Figure 5:**
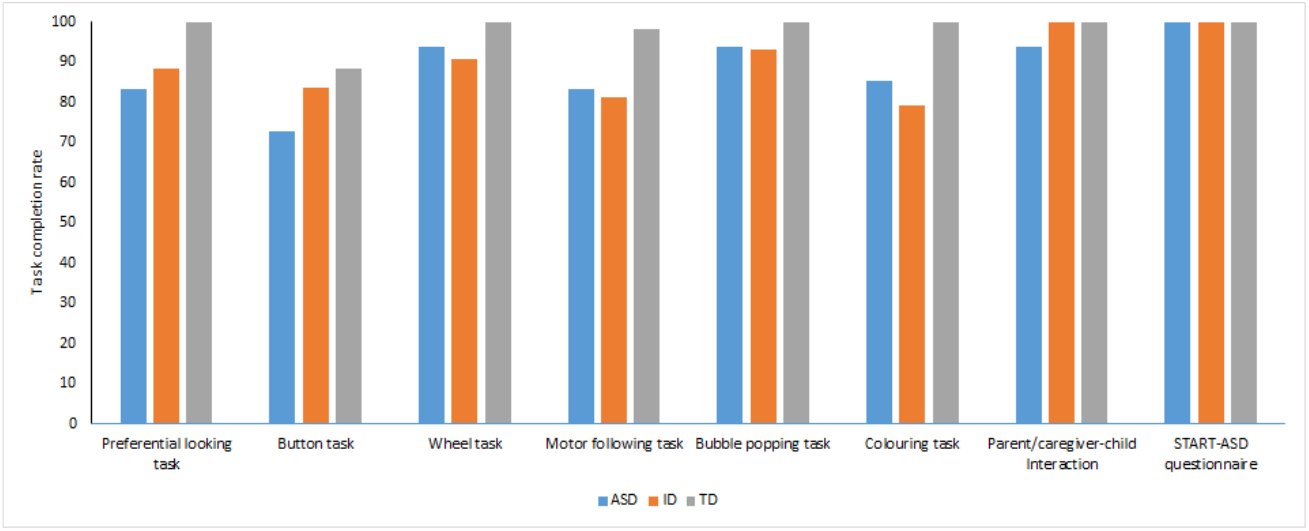
Task completion rate for each START task for each of the three groups of children (ASD, ID and TD)

Triangulation of data from the observation schedule and in-depth interviews (see Supplementary Table 4) with non-specialist health workers revealed that although conducting a child developmental assessment in households posed unique challenges such as limitations of space, variations in lighting and background noise and interruptions by family members, the health workers were able to identify suitable conditions to optimise data capture, particularly through cooperation with family members. Health workers specified that they found the detailed written standard operating protocols and scripts provided to them during training useful in guiding assessments. Interestingly, while some design elements in the app facilitated its smooth administration, other innovations, such as webcam video-based eye-tracking, led to confidentiality concerns in parents. App-based assessment seems to have high acceptability for children, who engaged well with the health workers, actively played the “games” on the tablet and enjoyed its child-friendly design elements. As expected, TD children were better able to engage with the team and the task than were the children with neurodevelopmental disorders, but parents of children with neurodevelopmental disorders reported that their child was comfortable during these assessments. Results also show that parents found START acceptable but questioned the credibility of an app-based assessment of child development.

## DISCUSSION

We tested a battery of tasks, questionnaires, and observational measures administered by a non-specialist on a mobile platform (app) in three groups of children with and without neurodevelopmental disorders. This mobile platform was found to be both feasible for delivery by non-specialists in a community-based setting in homes and acceptable to all users including community health workers, parents and children. Workers were able to collect quantitative, multidimensional data in cooperation with families; this experience is consonant with those of others working with profoundly autistic children (McKinney et al., 2021). We find strong evidence for group differences on the majority of the measures between children with and without neurodevelopmental disorders, and comparatively little evidence for differences between the two groups of children with neurodevelopmental disorders (ASD and ID).

### Task measures

The task measures focused on social, sensory, and motor functioning. Significant group differences between the TD and neurodevelopmental disorder groups were noted in tasks within each of these three domains. Specifically in the social domain, greater attention to social over non-social rewards was noted in TD children, using a preferential looking task. This pattern of results is consistent with reports on similar paradigms applied in laboratory settings, using standard infra-red eye trackers (Hedger et al., 2020). This observation supports the view that preferential looking toward social stimuli can be a proxy for social features of autism, and is not a proxy for general cognitive ability. It is also consistent with the results of a recent meta-analysis that showed no effect of IQ on preferential looking toward social stimuli (Hedger et al., 2020). Reduced attention to social stimuli has been widely noted in autistic children and adults, and has been suggested to be predictive of autistic symptomatology in later childhood (Bacon et al., 2020).

In contrast to the preferential looking task, where participants view the stimuli passively, the button task involves making an instrumental choice. Contrary to expectations, the button task did not show a difference between the three groups. This absence of any group difference could be driven by differences in the administration of the task between the current and the original report on this paradigm (Ruta et al., 2017). In the original report, the contingencies between the button and the associated stimuli (social/non-social) were not explicitly told to the child. In contrast, the current administration involved the child being clearly shown how one of the buttons was associated with the video of a child while the other was associated with a set of moving shapes. This change might have reduced the child’s motivation to explore on his/her own, leading to roughly equal numbers of button presses for the social or the non-social stimulus. It also is possible that this apparent inconsistency might be driven by cultural and/or ascertainment-related differences in this sample from a tertiary clinic in urban India, in contrast to those of the Italian sample on which the first report was based.

Strong group differences were noted in all task measures of motor function. The TD group performed more accurately than both the ASD and ID groups in the motor following task, as indexed by lower spatial errors (RMSE). However, no group differences were noted in the jerk profile in this dynamic visuomotor task, as have previously been shown in adults with autism using a gross motor task (Cook et al., 2013) and in autistic children using a visuomotor task of touchscreen movements to static targets (Weisblatt et al., 2019). Convergent findings indicating poorer visuomotor control in autistic children compared to the TD group were demonstrated as greater numbers of boundary crossings in the colouring task, and lower accuracy in reaching a dynamic target in the bubble-popping task. Additionally the ASD group used significantly greater force than the TD group in this task, replicating similar results reported using a different tablet-based task in children with ASD (Anzulewicz et al., 2016). Greater force in hitting a target on the tablet as well as spatial targeting errors could be interpreted as a manifestation of poor closed-loop motor control, wherein autistic users are less able to decelerate their action on time to hit a precise location on the tablet screen. Poor motor control can result from a reduced use of sensory information to adjust motor behaviour as an online process (Haswell et al., 2009), and is consistent with longstanding theoretical models of sensorimotor and cognitive prediction error in autism (Courchesne & Allen, 1997; Van de Cruys et al., 2014; Sinha et al., 2014; Palmer et al., 2017).

In the domain of sensory sensitivity, we used a tablet adaptation of a task previously associated with group differences between children with and without ASD (Tavassoli et al., 2016). Greater preference for specific types of sensory stimuli such as spinning wheels has been widely observed in autism (Sasson et al., 2011). While the underlying mechanisms for enhanced interest in such stimuli remain poorly understood, one feature shared by these stimuli is high predictability. We observed an expected pattern of group differences in the current sample, with autistic children spending longer time looking at the spinning wheel with illusory contours compared to the TD children. As such, this result demonstrates generalisability of the original observations with a real spinning wheel, and points to the potential for a scalable assay of visual sensory sensitivity.

### Parent/Caregiver-Report and Interaction measures

The parent/caregiver-report questionnaire was based closely on a tool specific for identification of ASD in an Indian context (INDT-ASD). Unsurprisingly, scores on this questionnaire significantly differed between all three groups (ASD, ID, TD) in the expected direction, replicating previous reports with the original tool (Gulati et al., 2019; Juneja et al., 2014).

The caregiver-child interactive episode revealed substantial atypicality in both key metrics of interaction. Autistic children initiated social interaction less than the TD group did, and also trended toward fewer initiations compared to the ID group. Fewer synchronous responses from the caregiver were evoked in interaction with ASD and ID children, compared to those with TD children. This result is consistent with an earlier report of reduced synchronous parent-child interactions in autistic relative to TD children (Feldman et al., 2014).

The majority of the START measures showed the expected pattern of group differences between autistic children and their TD counterparts in home settings. These data demonstrate a) the feasibility of administering a multi-domain assessment of autism-relevant phenotypic dimensions at home by non-specialist health workers, and b) the potential for scalability of this platform to other low-resource settings. Two caveats do need to be noted. First, there was little evidence for specificity of these measures in discriminating between the ASD and ID groups. To investigate this apparent equivalence further, we re-examined each case’s clinical notes, which revealed two insights: a) all of the ASD cases also met criteria for ID. This observation reflects the ground realities in India, where most ASD diagnoses in children within tertiary centres are at the severe end of the spectrum, and likely to be associated with developmental delay. b) A majority of the children in the ID group showed significantly elevated autistic symptoms. The phenotypic overlap in these groups could therefore have contributed to the observed lack of group differences between ASD and ID children. Second, all children in the atypical neurodevelopmental groups came from a tertiary clinic, which would have introduced a bias toward more severe levels of dysfunction, leading to greater group differences and also, as aforementioned, potentially to inconsistencies with previous results such as the absence of group difference on the instrumental social choice button task.

These caveats notwithstanding, we combined all the measures to test their ability to discriminate the ASD, ID, and the TD groups using machine-learning. This analysis revealed an overall accuracy of 78% in classifying the three groups, a considerable boost from the accuracy achieved by any of the measures alone. This result highlights the advantages of a multi-measure platform that combines task performance with parent/caregiver-report to achieve greater precision in assessing autism.

## Conclusion

The current study demonstrates the potential and proof of principle for a tablet-based app for assessing autistic children that can be administered by non-specialist health workers with minimal training. The app includes task, questionnaire, and observational assessments of aspects of behaviour that index social, sensory, and motor function. Individual metrics from each task showed a consistent pattern of differences between typically and atypically developing children. Combining the information from multiple measures within the app resulted in high classification accuracy for the three groups of children (ASD, ID, TD). Future work should test this app prospectively in a large population-based study to assess the predictive validity of each of these measures independently, and in combination, for atypical neurodevelopmental status. The eventual goal for this platform is its incorporation into a routine health surveillance system to help identify children with- or at risk of- neurodevelopmental disorders.

## Data Availability

All data will be made available upon request from the corresponding author following the publication of the article in a peer-reviewed journal.

## Acknowledgements

This work was funded by a Medical Research Council Global Challenge Research Fund grant to the START consortium (PI: Chakrabarti; Grant ID: MR/P023894/1). The authors express their gratitude to Geraldine Dawson for sharing the protocol for parent-child interaction, to Therapy Box UK for app development, and to all families and health workers who took part in this study.

## Footnotes

1 We recognise that the autism community has a diversity of views in using person-first terminology. To reflect this diversity of views, we use ‘children with ASD’ interchangeably with ‘autistic children’ throughout the manuscript.

