## Supplementary Material for "Using mobile health technology to assess childhood autism in low-resource community settings in India : an innovation to address the detection gap"

| | | |
|---|---|---|
| Section1.1 Dependent variable calculation for the motor following task | page | 2 |
| Section 1.2 Machine learning data analysis | page | 3 |
| Supplementary Table 1: Results from Machine Learning analysis | page | 4 |
| Supplementary Table 2: Alternative test statistics for group differences | page | 5 |
| Supplementary Table 3: Alternative test statistics for group differences | page | 7 |
| Supplementary Table 4: Summary of qualitative results | page | 8 |
| Section 1.3 START- ASD questionnaire | page | 10 |
| Section 1.4 Observation Schedule | page | 12 |
| Section 1.5 Interview Schedule for caregivers/families | page | 14 |

#### 1.1 Motor following task:

Root Mean Square Error (RMSE) in the motor following task was calculated using the following formula

$$RMSE = \sqrt{\sum \frac{(x_{pred} - x_{ref})^2 + (y_{pred} - y_{ref})^2}{N}}$$

where $x_{pred}$ and $y_{pred}$ are the participant's finger position on x and y axes on the screen while $x_{ref}$ and $y_{ref}$ are the corresponding positions for the butterfly. $N$ indicates the total number of recorded data points for a test attempt.

Additionally, we analysed the 'frequency gain' metric for all participants using a Fast Fourier Transformation (FFT). For this the trajectories of the source and target motion are resolved into multiple waves of varying amplitudes using FFT. This allows us to analyze the closeness in the source and target motions along both axes by observing them in the frequency domain. This is achieved by calculating the average gain in amplitude for the source motion in the vicinity of each target frequency. This gain represents the accuracy of the source in copying the target trajectory, where the resultant value approaches one in case of high degree of correspondence between the two trajectories. More specifically, $G_f$ the gain at a given frequency $f$, is calculated as

$$G_f = \frac{U_{fm}}{B_f}$$

Here, $B_f$ is the amplitude for target's (butterfly) motion at frequency $f$; and $U_{fm}$ is the average amplitude of user's (source) motion in the vicinity of frequency $f$ (a neighborhood of three frequency bins including $f$ is used). It is given as

$$U_{fm} = \frac{U_{f_{-1}} + U_f + U_{f_{+1}}}{3}$$

Here, $U_f$ represents the amplitude of user's motion at frequency $f$. The subscripts $f_{-1}$ and $f_{+1}$ represent frequencies adjacent to frequency $f$. Target motion is pre-determined and hence its amplitude is not approximated. The child is assumed to be following this motion and hence $U_f$ is approximated at the central bin at frequency $f$.

In addition, Jerk, the change in acceleration per time, was derived as the third-order differential of the participant's trajectory with respect to time.

### 1.2 Machine learning-based data analysis:

As per current best practices, the machine learns from a subset of the labelled data and creates a classification model; the model is then subjected to the remaining unseen data to determine the desired accuracy. Classification accuracies have been reported for a specific group as:

$$C_{ASD} = \frac{P_{ASD}}{N_{ASD}}$$

where, $P_{ASD}$ is the number of children correctly classified as ASD, $N_{ASD}$ is the actual number of ASD children, and thus the ratio $C_{ASD}$ is the classification accuracy for the ASD group (reported in %). $C_{ID}$ and $C_{TD}$ have been similarly derived from the pairs $P_{ID}$, $N_{ID}$ and $P_{TD}$, $N_{TD}$, respectively. Finally, the overall classification accuracy $C_{Overall}$ is determined as:

$$C_{Overall} = \frac{P_{ASD} + P_{ID} + P_{TD}}{N_{ASD} + N_{ID} + N_{TD}}$$

A 5-fold cross-validation scheme has been followed to minimize any bias and variance which might be introduced due to the relatively small size of our dataset (a deep neural net-based model may not be feasible).

**Supplementary Figure 1:** Analyzing all possible combinations of features for classification accuracy in a 100 round polled scheme

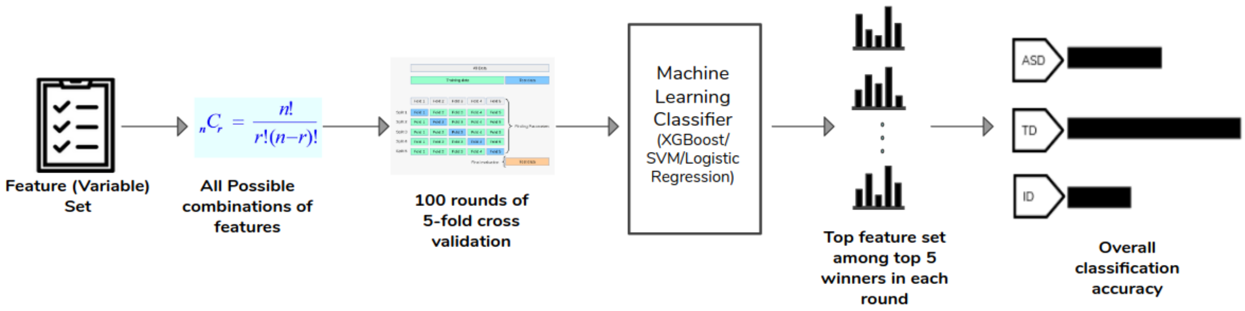

To determine the set of features from the combinatorial set (all possible combinations of features were evaluated) with the best overall accuracy, a polling scheme consisting of 100 rounds was used as shown in supplementary Figure 1. In each round, a 5-fold cross-validation experiment was performed for all possible sets or combinations of features. The top 5 winning combinations of features, based on the overall classification accuracy, were noted for each round. Finally, the most frequently occurring feature combination was declared as the overall winner and its average accuracy, across the 100 rounds, is reported. This polling scheme ameliorates any undesirable effect of outliers and the cherry picking of favourable results.

**Supplementary Table 1:** Results from Machine Learning analysis. Overall classification accuracy for each feature vector (dependent variable) is listed.

| Task | Dependent Variable ID | Dependent Variable | Mean Overall Classification Accuracy (%) | Mean proportion % of subjects across different groups (ASD:ID:TD) |
|---|---|---|---|---|
| **Preferential Looking Task** | (1) | Social preference | 42.75 | 34:32:34 |
| **Button Task** | (2) | Social choice | 46.80 | 29:35:36 |
| **Wheel Task** | (3) | Proportion of looking at the wheel, Minimum distance, Maximum distance | 52.22 | 36:33:31 |
| **Motor Following Task** | (4.1) | RMSE | 60.01 | 34:32:34 |
| | (4.2) | FFT X Axis, FFT Y Axis | 49.22 | 32:31:37 |
| | (4.3) | Jerk | 41.09 | 34:31:35 |
| **Bubble Popping Task** | (5.1) | Distance on X Axis, Distance on Y Axis | 49.04 | 34:33:33 |

| | | | | |
|---|---|---|---|---|
| | (5.2) | Force | 52.33 | 34:33:33 |
| **Colouring Task** | (6.1) | Crossing over | 48.34 | 30:30:40 |
| | (6.2) | Proportion of color spilt out to the total area inside figure | 49.18 | 33:30:37 |
| **Parent/Caregiver- Child Interaction** | (7) | Caretaker: Synchronous response, Child: Initiation | 50.19 | 34:33:33 |
| **Questionnaire** | (8) | Total Score | 60.11 | 37:33:30 |

**Supplementary Table 2**: Alternative test statistics when assumption of homogeneity of variance is violated

| Task | Variable | Levene's test p value | Robust test: Welch | Robust test: Brown-Forsyth | Post-hoc contrasts (Games-Howell), p-value |
|---|---|---|---|---|---|
| **Domain: Social** | | | | | |
| **Preferential looking task** | Social looking | .216 | NA | NA | NA |
| **Button task** | Social preference | .145 | NA | NA | NA |
| **Domain: Sensory** | | | | | |
| **Wheel task** | Duration | .179 | NA | NA | Na |
| | Maximum distance | .002 | 6.64, $p = .002$ | 7.58, $p = .001$ | TD < ASD, .002<br>TD ~ ID, .183<br>ASD ~ ID, .080 |
| | Minimum distance | .049 | 20.20, $p <.0001$ | 17.73, $p <.0001$ | TD < ASD, <.0001<br>TD < ID, <.0001<br>ASD ~ ID, .629 |

| | | | | | |
|---|---|---|---|---|---|
| **Domain: Motor** | | | | | |
| **Motor following task** | RMSE | <.0001 | 41.08, $p$ <.0001 | 32.86, $p$ <.0001 | TD < ASD, <.0001<br>TD < ID, <.0001<br>ASD > ID, .005 |
| | FFTx | .024 | 9.64, $p$ <.0001 | 6.92, $p$ = .002 | TD < ASD, .004<br>TD < ID, .003<br>ASD ~ ID, .999 |
| | FFTy | .002 | 16.40, $p$ <.0001 | 14.55, $p$ <.0001 | TD < ASD, <.0001<br>TD < ID, <.0001<br>ASD ~ ID, .277 |
| | Jerk | .749 | NA | NA | NA |
| **Bubble popping task** | Force | .050 | 9.57, $p$ <.0001 | 8.50, $p$ <.0001 | TD < ASD, <.0001<br>TD ~ ID, .086<br>ASD ~ ID, .152 |
| | Distance X | <.0001 | 23.23, $p$ <.0001 | 14.93, $p$ <.0001 | TD < ASD, <.0001<br>TD < ID, <.0001<br>ASD ~ ID, .065 |
| | Distance Y | <.001 | 17.33, $p$ <.0001 | 11.97, $p$ <.0001 | TD < ASD, <.0001<br>TD < ID, <.0001<br>ASD ~ ID, .283 |
| **Colouring task** | Crossover errors | .021 | 19.99, $p$ <.0001 | 15.60, $p$ <.0001 | TD < ASD, <.0001<br>TD < ID, <.0001<br>ASD ~ ID, .672 |
| **Domain: Parent report/observation** | | | | | |
| **Parent/ Caregiver-Child Interaction** | Caretaker's synchronous interaction | .001 | 9.21, $p$ <.0001 | 11.15, $p$ <.0001 | TD < ASD, <.0001<br>TD < ID, .013<br>ASD ~ ID, .246 |
| | Child's initiation | .624 | NA | NA | NA |
| **Questionnaire** | Sum score | .006 | 56.82, $p$ <.0001 | 45.71, $p$ <.0001 | TD < ASD, <.0001<br>TD < ID, <.0001<br>ASD > ID, <.0001 |

**Supplementary Table 3**: Kruskal-Wallis test for group comparison for tasks where assumption of normality is violated

| **Task** | **Variable** | **Kolmogorov-Smirnov test** | **$X^2$** | **df** | ***p*** | **$\mathcal{E}^2$** | **Dwass-Steel-Critchlow-Flinger pairwise contrasts, p-value** |
|---|---|---|---|---|---|---|---|
| **Domain: Social** | | | | | | | |
| **Preferential looking task** | Social looking | 0.06, *p* =.73 | NA | NA | NA | NA | NA |
| **Button task** | Social preference | 0.11, *p* = 0.16 | NA | NA | NA | NA | NA |
| **Domain: Sensory** | | | | | | | |
| **Wheel task** | Proportion duration of watching | 0.19, *p* <.001 | 10.35 | 2 | 0.006 | 0.09 | TD < ASD, 0.006<br>TD > ID, 0.047<br>ASD ~ ID, 0.773 |
| **Domain: Motor** | | | | | | | |
| **Motor following task** | RMSE | 0.10, *p* = 0.167 | NA | NA | NA | NA | NA |
| | FFTx | 0.09, *p* = 0.403 | NA | NA | NA | NA | NA |
| | FFTy | 0.09, *p* = 0.278 | NA | NA | NA | NA | NA |
| | Jerk | 0.36, *p* <.001 | 6.83 | 2 | 0.033 | 0.06 | TD > ASD, 0.050<br>TD ~ ID, 0.139<br>ASD ~ ID, 0.548 |
| **Bubble popping task** | Force | 0.09, *p* = 0.335 | NA | NA | NA | NA | NA |
| | Distance X | 0.17, *p* = 0.002 | 37.46 | 2 | <.0001 | 0.31 | TD < ASD, <.001<br>TD < ID, <.001<br>ASD ~ ID, 0.316 |

| | | | | | | | |
|---|---|---|---|---|---|---|---|
| | Distance Y | 0.13, *p* = 0.031 | 25.76 | 2 | <.0001 | 0.22 | TD < ASD, <.001<br>TD < ID, <.001<br>ASD ~ ID, 0.869 |
| **Colouring task** | Crossover errors | 0.10, *p* = 0.277 | NA | NA | NA | NA | NA |
| **Domain: Parent report/observation** | | | | | | | |
| **Parent/ Caregiver-Child Interaction** | Caretaker's synchronous interaction | 0.09, *p* = 0.394 | NA | NA | NA | NA | NA |
| | Child's initiation | 0.11, *p* = 0.189 | NA | NA | NA | NA | NA |
| **Questionna-ire** | Sum score | 0.17, *p* = 0.001 | 61.23 | 2 | <.0001 | 0.47 | TD < ASD, <.001<br>TD < ID, <.001<br>ASD > ID, <.001 |

**Supplementary Table 4**

Summary of themes and subthemes emerging from the interview of health workers and parents.

| Topic | Theme, sub-theme, quotation |
|---|---|
| Health worker's experience using the START app | Facilitators to smooth administration: Statement of Procedure, script, and app design elements<br><br>*We have been given words [script]– if we speak them as it is, we remain confident.* (health worker 1)<br><br>*When the game finishes, a small dialogue box appears on the screen that this game is finished and we press the arrow button to go to the next game. It helps a lot. We get to know that have to go to next [game].* (health worker 2)<br><br>*If a child didn't take interest in wheel task, we switched to button or butterfly task and so on…and if a child is not at all interested in playing on the tablet then we used to record PCI. [we would say] "It's fine if you don't want to play on a tablet. See! We have got toys for you, let's play with them".* (health worker 1) |
| | Suitability of household environment for data capture |

| | |
|---|---|
| Challenges faced by health workers during data collection and strategies adopted to overcome them | Sub-theme 1: Availability of space in households<br><br>*Some families had a single room house – they were living and eating in the same room. In these cases, adult family members used to go out while we made siblings sit in a corner.* (health worker 1) |
| | Sub-theme 2: Disruptions by family members<br><br>*We ensured that ... no other family members except mother-child are in the room.* (health worker 2) |
| | Sub-theme 3: Disruptions in the testing environment<br><br>*If an air cooler was on then we requested them (parents) to switch it off or if a phone was ringing in the room then we gestured to them to put it on silent mode.* (health worker 2) |
| | Engaging atypical children<br><br>*There is a difference between a normal child and child with problems. A normal child engages with us quickly but a child with problems might not be comfortable in sitting with us.* (health worker 2) |
| | Confidentiality concerns<br><br>*Consenting video has really helped in giving a clear picture to the families (about the assessment)...Also, families had concerns – will these games cause any harm to the child and will the video be uploaded to any website or shown publicly.* (health worker 2) |
| Acceptability to children | Interest in digital devices<br><br>*Nowadays children like laptops or tablets if you make them play on it, they like it. It could be any game.* (Father, ID child)<br><br>*He was interested and accordingly the assessment proceeded smoothly.* (Father, ASD child) |
| | App design elements<br><br>*It (START task battery) was appropriate for them. Otherwise the child gets bored and runs away.* (Mother, ASD child)<br><br>*He liked bursting bubbles and colouring* (Father, ASD child) |
| | Health worker engagement<br><br>*She (health worker) was able to understand how to deal with the child.* (Mother, ASD child)<br><br>*The health worker was doing it nicely – she was explaining to the child quite well.* (Mother, ID child) |
| Acceptability to parents | Overall high acceptability of the app |

| | |
|---|---|
| | *It was nice but she (child) wasn't so successful in games (wasn't able to play well). She is quite young so accordingly it was fine.* (Mother, ID child) |
| | Skepticism of apps as a valid assessment of child development<br><br>*Suppose any child has been identified and a highly qualified doctor from your team explains it to them then they would feel that their child actually requires it (intervention)...How will they get convinced through an app? Obviously, they would need a doctor.* (Father, ASD child). |

#### 1.3 START-ASD Questionnaire

Instructions for the health workers: Please read out the items to the caregiver and ask them to choose from the options.

| No | Items in English | Option |
|---|---|---|
| 1(R) | Does your child look at your face or eyes, when you talk to him/her? | Yes/no |
| 2(R) | Does your child engage in pretend play, such as using something like an imaginative phone by putting it on ear and talking, pretending to cook using toy utensils, making sound of a car/auto/bike/rail while moving something etc. | Yes/no |
| 3(R) | Does your child play cooperatively with other children or with you? Like throwing ball, hide and seek, peek-a-boo etc. | Yes/no |
| 4 | Does your child get disturbed by usual sound or light? Such as getting annoyed by the sound of the kitchen utensils and trying to close the ears with hands/fingers, not able to bear the sound of the vehicles, unable to bear the fairy/festival lights, gets irritable by the sharp light of the bulb, etc. (Social worker please ask the opposite behaviour too, such as does the child like loud sounds or sharp lights? He/she watches bright lights by going close to them and/or listen to the radio / TV by sticking ears to them?) | Yes/no |
| 5(R) | Does your child imitate you? Like making gesture for "bye-bye" or hello, or wearing a scarf or bag like you? | Yes/no |

| | | |
|---|---|---|
| 6 | Does your child get annoyed with cloth tags, woollen or tight cloths, toothbrushes, socks etc. Or does he like rubbing some items / cloth on his body repeatedly even if it results in scratches. | Yes/no |
| 7(R) | Is your child able to use language according to his/her age? Like adding words to make sentence "let's go out", or to answer you correctly and asking questions "what is that?", "when are we going?" etc. | Yes/no |
| 8 | Does your child call himself by his/her name like "Vivek will eat food". | Yes/no |
| 9(R) | Does your child show you the things he/she likes by pointing fingers to them? | Yes/no |
| 10 | Does your child repeat any kind of movement frequently? Like constantly making flapping/wriggling movement with his hands/fingers, constantly moving the body back and forth while sitting, constantly moving the head or body in unusual manner, etc. | Yes/no |
| 11(R) | Does your child look at you / responds when called by name? | Yes/no |
| 12 | Does your child repeat certain voices, such as the sharp (high pitched) meaningless sounds, repeating your spoken words without context or meaning, repeating any sound heard on TV/redio/computer meaninglessly? | Yes/no |
| 13(R) | Does your child come to you and show you when he/she has done something good? | Yes/no |
| 14 | Does your child play oddly with toys? Such as instead of using them meaningfully he/she just lines them up, or instead of running the toy car he spends long time looking at its wheels, smells or rubs toys on his body. | Yes/no |

**Scoring:** The items with the (R) indicate reverse scoring i.e. a score of 1 is given for “No” and for the other items a score of 1 is given for “Yes”. Then the sum is calculated to get the severity of autistic symptoms.

**1.4 Observation schedule used by the research assistant**

Child code
Date of assessment
General observations
Specific observations
*Observation coding*

| | *Low* | *Medium* | *High* |
|---|---|---|---|
| *Mother factors* | | | |
| Interest in the visit | 1 | 2 | 3 |
| Favourable reaction to tablet | 1 | 2 | 3 |
| Distractions with other duties | 1 | 2 | 3 |
| Distractions with other family members | 1 | 2 | 3 |
| *Child factors* | | | |
| Exposure to smartphone/tablet | 1 | 2 | 3 |
| Child's interest in the assessment | 1 | 2 | 3 |
| Did parent have to help child engage with the assessment | | | (Y/N) |
| Ability to swipe | 1 | 2 | 3 |
| Ability to tap | 1 | 2 | 3 |

Total time of engagement (establishment of rapport with the child) Minutes

| | *Low* | *Medium* | *High* |
|---|---|---|---|
| *Environment factors* | | | |
| List all family members present (other than mother/father) | | | |
| Number of times siblings disrupted assessment | | | times |
| Number of times other family members disrupted assessments | | | times |
| Any other types of disruption to assessment | | | times |
| Interest of other family members in the assessment | 1 | 2 | 3 |
| Level of noise in assessment room | 1 | 2 | 3 |
| Level of light in assessment room | 1 | 2 | 3 |

Type of lighting Natural Artificial (Bulb) Artificial (Tubelight) Torch

| *Assessor factors* | *Low* | *Medium* | *High* |
|---|---|---|---|
| How well was assessment process explained to mother | 1 | 2 | 3 |
| How well was the child engaged by assessor | 1 | 2 | 3 |
| How well did the assessor judge mood of the child | 1 | 2 | 3 |
| How well did the assessor administer the devices | 1 | 2 | 3 |
| How well did the assessor manage the family | 1 | 2 | 3 |

| Eye tracking | *Low* | *Medium* | *High* |
|---|---|---|---|
| What was the arrangement | | | |

table/chair floor/table floor/chair 2 chairs bed/chair others (specify)

| | | | |
|---|---|---|---|
| How difficult was it to get a suitable arrangement | 1 | 2 | 3 |

Did the child need to sit in the mother's lap (Y/N)
Was mother's face coming in the parameters screen (Y/N)
Was mother prompting the child during the assessment (Y/N)
Did the child try to touch the tablet during eye tracking (Y/N)
Was there a need to move to another task and then back (Y/N)
Time taken to calibrate the parameters immediately within 3 minutes not at all
Did the child disengage from the task (Y/N)
Was the task aborted (Y/N)
Mention reason:

Wheel task
No of demos needed 1 2 3
Did the child understand the task (Y/N)
Did the child lose interest during play mode (Y/N)
Was the task aborted (Y/N)
Mention reason:

Button task
No of demos needed 1 2 3
Did the assessor have to hold the child's hand during demo (Y/N)
Did the child understand the task (Y/N)
Did the child lose interest during play mode (Y/N)
Was the task aborted (Y/N)
Mention reason:

Butterfly task
No of demos needed 1 2 3
Did the assessor have to hold the child's hand during demo (Y/N)
Did the child understand the task (Y/N)
Did the child lose interest during play mode (Y/N)
Was the task aborted (Y/N)
Mention reason:
Bubbles task
No of demos needed 1 2 3
Did the assessor have to hold the child's hand during demo (Y/N)
Did the child understand the task (Y/N)
Did the child lose interest during play mode (Y/N)
Was the task aborted (Y/N)
Mention reason:
Colouring
No of demos needed 1 2 3
Did the assessor have to hold the child's hand during demo (Y/N)
Did the child understand the task (Y/N)
Did the child lose interest during play mode (Y/N)
Was the child pressing too hard i.e. colour not coming (Y/N)
Was the task aborted (Y/N)
Mention reason:
PCI observations
Questionnaire observations

**1.5 Interview schedule used to evaluate acceptability of the assessment from caregivers/families**

The purpose of the in-depth interview (IDI) with mothers of children completing the START assessment was to understand the acceptability and feasibility of using START in Delhi households. Permission to audio record the interview was taken prior to the interview. If the parent was uncomfortable with audio recording, permission for note taking during the interview was sought. The following information was provided to the mother to guide the interview process:

"Thank you for meeting me today and for participating in our study. We had visited you at your home to carry out a tablet assessment that we are developing. I would like to understand more about your experience of the assessment by asking you a few questions. I am interested in knowing *your opinions*/suggestions and you can refuse to answer any question in case you feel uncomfortable. Could we begin?

*Experience of the consenting process*

You were approached by a health worker who explained the purpose of the study and requested for a time when she could visit you at home.

- Could you describe how you felt when you were approached by the health worker?
- Could you describe any immediate concerns you had about the assessment/home visit?

*Experience during the visit*

I would like to know more about your experience during our visit.

What did you like about the assessment? What did you dislike?

Probes: Time duration of the visit, comfort with a tablet assessment, comfort with video recording.

What did your family think about the assessment?

*Child engagement with START:*

What was your child's reaction to the health worker visiting them and during the assessment?

Probe: what do you think might be the reasons that he/she enjoyed/did not enjoy our visit?

What are your suggestions to make this more enjoyable for other children in the future?

*Health worker training:*

What did you think of the way the tablet was administered?

Would you have liked the health worker to do anything differently?

Probe: Was the health worker sensitive to your child's requirements/needs during the assessment?

Could you describe any concerns you had during the assessment process? How did the health worker address these?

*Scaling up*

In the future, we would like to carry out this assessment with more children at their homes.

How would other families like yours respond?

What are your suggestions so that most families would be happy to participate?

I would like to thank you on behalf of our team for taking the time out to not only be a part of our work but also for speaking with me today. Your feedback is very important for us to understand how we can make this experience better for families with young children."
